# Analysis of requests for reservation of blood concentrates in patients referred to urgent surgeries in a teaching hospital

**DOI:** 10.1101/2023.01.30.23285131

**Authors:** Cintia de Lelis Garcia, Michelle Petrolli Silveira, Seleno Glauber de Jesus-Silva

## Abstract

**Introduction:** The rates of request and use of blood concentrates are still poorly reported in the literature. This study aimed to analyze the rates of requests for blood concentrates and their use in patients undergoing emergency surgery in a teaching hospital.

**Methods:** A retrospective, quantitative and descriptive study was conducted in 359 medical records of patients in urgent surgery scheduled with a request for a reserve of blood concentrate. The ratios between crossmatched and transfused units (C/T), transfusion index (TI), and probability (TP) were calculated, and the times between request and delivery at the transfusion agency (TA) and patient admission to the surgical center (SC).

**Results:** The mean age was 58.5 ± 22.2 years, with the majority being male (53.1%). There was an average of 27.5 monthly requests (min 12, max 44). Ninety-seven units of blood concentrates were transfused into 44 patients (C/T ratio 7.59; TI 0.27; TP 12.3%). Only seven patients had their requests made after admission to the OR. The median time between the request and arrival at the TA was 1h15min, while that between the request and the patient’s arrival at the SC was 5h23min. There was greater transfusion in major surgery (major 37, 14.8% vs. medium 7, 6.5%; p = 0.027) and non-orthopedic surgery (orthopedic 9, 4.0% vs. non-orthopedic 35, 26.9%; p < 0.001).

**Conclusion:** there was a significant discrepancy between the number of requests for blood reservation and its real use and an increased time between reservation requests and their arrival at the TA.

## Introduction

In the routine of in-hospital elective surgeries, standard operating protocols are developed to organize the flow and maintain patient safety. According to the World Health Organization (WHO), surgical patient safety consists of the surgical team recognizing and preparing for potential blood loss. It is recommended that a team member check the availability of blood components before anesthetic induction^1^.

With the increase in demand for elective surgeries, the excess of blood component reserve requests and the number of unused bags have become a constant. With the increase in longevity and high-risk interventions, a greater demand for blood components for older patients has also been observed^2^.

The reserve of blood components follows a pre-established flow, depending on the type of surgery. The transfusion agency (TA) and the nursing team are informed about collecting blood samples for blood typing, investigating irregular antibodies, and crossmatching. However, when there is no previous blood typing, a low supply of rare blood groups, or intercurrences in patients without prior reservation, there may be a delay in the release of the blood components^3^. The blood reserve must be done in advance, filling out a standardized form and the medical prescription. All documents and the identified patient’s blood sample must be forwarded up to 48 h in advance, according to the institution’s protocol. This allows time for sample testing, proof of compatibility, and verification of available stock^4^.

Based on the institutional protocol, blood reservations must be made consciously, as prepared, and non-transfused blood components lead to greater material consumption, more significant financial expense, and service time. The blood component units prepared as a surgical reserve for a patient will not be available for other patients; the surgeon confirms that the reserved blood components will not be used intraoperatively or in the immediate postoperative period, which may lead to the product being discarded^5^. Additionally, the response time between the reservation request and the actual transfusion in the patient is a measure of efficiency^6^.

Despite being a critical topic in the hospital dynamics, those parameters are still little reported in the scientific literature. There is a scarcity, not only at the national level, of data on the dynamics of blood component requisition and their usage rates. Considering the demand for urgent surgeries in large hospitals, the need to use standard operating protocols (SOP) for the reserve of blood components for surgeries, and the scarcity of loco-regional data, this study aimed to analyze the transfusion and request rates for blood concentrates and their use in patients undergoing emergency surgery at a teaching hospital.

## Material and methods

### Design of the study

This was a cross-sectional study conducted by analyzing 359 medical records of patients who were referred to the surgical center (SC) to undergo urgent surgery and who had a request for a reserve of blood concentrates, from March 2020 to March 2021. The hospital is an “open door” unit with a population coverage of 300 thousand inhabitants. Furthermore, it is a quaternary hospital that performs solid organ transplants and is a reference in high complexity in cardiovascular surgery.

### Sample size

Based on an estimate that at least 37% of the surgeries referred to the SC did not follow the reservation protocol within the pre-defined period^7^, using a confidence level of 95% and an estimated error of 5%, for an infinite population, the minimum sample value calculated was 348 surgeries. Sampling was performed sequentially, not probabilistic, in order of entry into the SC from the start date of the investigation (March 2020), according to the administrative records of the sector.

### Data collection

Urgent surgeries were defined as those required to be performed within 48 h of hospital admission. Elective surgeries were considered in all those whose execution could await an appropriate circumstance and be rescheduled if necessary. Emergency surgeries were those that required immediate surgical treatment at risk of death.

Data collection from the medical records (physical and electronic) of the patients included in the study was performed by checking requests for blood concentrate reservations sent to the institution TA. Patients of both sexes, of any age, who underwent urgent surgical treatment, with a request for a reserve of blood concentrate at any time of hospitalization, either through the Brazilian universal public health system (*Sistema Único de Saúde*, SUS), health plans or private hospitalizations, between the predetermined months, were included. Exclusion criteria were medical records that did not contain basic information on whether to request blood transfusions, patient reoperations (same hospitalization), and patients with voluntary restriction of blood transfusion (Jehovah’s Witnesses). Elective and emergency surgeries were excluded from the study.

To describe the profile of the study participants, primary clinical-epidemiological data (sex, age, comorbidities, indication for surgery, surgical size, and type of surgery performed) were collected, as well as data on the reservation request: the time of request and prescription, time of admission to the SC, performance or not of the proposed surgery, time of infusion of the blood product and completion or not of the free and informed consent form for a blood transfusion by the patient or companion.

Comorbidities were defined as described in the medical records and surgery notes. Heart disease was defined as any functional or structural disease of the heart, and central neuropathy as any neurological disorder that affected central functions. Chronic kidney disease was defined as admission serum creatinine > 1.5 mg/dL. Smoking and alcohol consumption were self-reported without detailing the number of packs/year or grams of alcohol/year.

### Outcomes

The information collected was entered into a data acquisition worksheet, and the following outcomes were obtained: sample distribution according to sex, mean age of patients transfused or not, occurrence and number of reservation requests and transfusions performed, mean time between the request for the blood concentrate reserve to the TA and the patient’s arrival at the SC. The ratio between the number of units subjected to the compatibility test (crossmatched) and the number of transfused units (C/T ratio) was calculated. A ratio lower than or equal to 2.5 is considered an indication of efficient use of blood^8^. Also, the probability of transfusion (PT), defined as the number of transfused patients divided by the number of typed patients (x100), and the transfusion index (TI), considered as the number of transfused units divided by the number of typed patients (values > 0.5 are considered effective use of blood)^8,9^, were calculated.

### Statistical analysis

Data were submitted to descriptive statistics, using relative and absolute frequencies, mean and standard deviation, or median and interquartile range, depending on the symmetrical or asymmetrical distribution of the values. Data normality was analyzed by its graphic distribution and by the Kolmogorov-Smirnov test. Identification of outliers was performed by the ROUT^10^ test. However, these values were included in the final analysis as they were considered correct. The comparison of groups with an asymmetric distribution was made by determining the 95% confidence intervals. The association of groups with normally distributed values was performed using the two-tailed unpaired Student’s t-test and groups with binary variables using Pearson’s chi-square test. An alpha value = 5% was considered. GraphPad Prism v. 9.0 (San Diego, CA, USA) was used.

### Ethical considerations

The study was approved by the Research Ethics Committee of the Faculty of Medicine of Itajubá (CAAE 40762220.9.0000.5559, decision nr. 4,515,089). All the study steps were conducted in accordance with the Declaration of Helsinki. As the study had a retrospective design based on the collection of information obtained from clinical records, under confidentiality and with respect to patients’ anonymity, the informed consent form for the research was waived.

## Results

The initially eligible sample consisted of 398 patient records, of which 40 were excluded due to the lack of information relevant to the study and after application of the exclusion criteria, totaling 10% of losses. Therefore, the final number of medical records evaluated was 358. Most of the patients studied were male (male: n = 190, 53.1% vs. female: n = 168, 46.9%). The mean age of the sample was 58.5 ± 22.2 years. Clinical data and types of the emergency surgeries analyzed are shown in Table 1. The average monthly request for blood concentrates for urgent surgeries was 27.5, ranging from 12 (April 2020) to 44 (October 2020). All patients or guardians signed the informed consent for the transfusion of blood components.

**Table 1.** Clinical data of the analyzed patients and types of urgent surgeries performed in the studied period (N = 358).

| Variable | n | % |
| --- | --- | --- |
| Age (years) (mean ± SD) | 58.5 ± 22.2 | |
| Sex |  |  |
| Male | 190 | 53.1 |
| Female | 168 | 46.9 |
| Comorbidities |  |  |
| SAH | 124 | 34.6 |
| DM | 54 | 15.1 |
| Heart disease | 41 | 11.5 |
| Central neuropathy | 21 | 5.9 |
| Smoking | 17 | 4.7 |
| Elitism | 11 | 3.1 |
| CKD | 6 | 1.7 |
| Types of surgery |  |  |
| Orthopedic | 228 | 63.7 |
| General | 68 | 19.0 |
| Neurosurgery | 21 | 5.9 |
| Gynecology | 14 | 3.9 |
| Vascular | 10 | 2.8 |
| Urological | 8 | 2.2 |
| Thoracic | 7 | 2.0 |
| Cardiac | 2 | 0.6 |
| Type of surgery |  |  |
| Major | 250 | 69.8 |
| Minor | 108 | 30.2 |

SD, standard deviation. SAH, systemic arterial hypertension. DM, diabetes mellitus. CKD, chronic kidney disease.

Forty-four patients were transfused, corresponding to 12.3% of the total requests. Of the 736 units of blood concentrates requested, only 97 were transfused, corresponding to a usage rate of 13.2% and a C/T ratio of 7.59. The reserve and transfusion indicators are shown in Table 2. A total of 343 patients with requested reserve were operated. The remaining 15 patients had their surgery suspended for various reasons: lack of availability of post-op ICU (3), death (2), limb edema (1), use of anticoagulants (1), hypotension (1), and unreported causes (7).

**Table 2.** Reserve and transfusion indicators for the 358 patients studied.

| Variable | value |
| --- | --- |
| Nr. of transfused patients | 44 |
| Transfusion Probability* | 12.3% |
| Nr. of requested blood bags | 736 |
| Nr. of transfused bags | 97 |
| C/T ratio | 7.59 |
| Transfusion index † | 0.27 |
| Nr. of patients whose reservation was made after arrival at the SC | 7 (2.0%) |
| Nr. of patients whose surgery was cancelled | 8 (2.2%) |

\*Number of transfused patients divided by the number of cross-matched patients. C/T ratio, number of units subjected to compatibility testing/number of units transfused. †Number of units transfused divided by the number of patients typed. SC, surgical center.

Table 3 shows the temporal indicators between the request for reservation and referral to the TA and the patient’s admission to the SC. It was noted that the median time between the request and its arrival at the TA was 1h15min, while the median time between the request and the patient’s arrival at the SC was 5h23min. Many requests (n = 104; 30.3%) were made within 6 h of the patient’s admission to the SC. In 44 patients (12.8%), the request for a reserve of blood concentrate was made more than 48 h after admission to the SC.

**Table 3.** Temporal indicators in relation to the request for a reserve of blood concentrate, its delivery at the transfusion agency and patient arrival at the surgical center.

| Variable | median | IQR | 95% CI |
| --- | --- | --- | --- |
| Time between request and delivery at TA (h:min) |  |  |  |
| All | 1:15 | 0:35 - 2:34 | 1:10 - 1:26 |
| Transfused | 0:50 | 0:18 - 1:32 | 0:26 - 1:08 |
| Not transfused | 1:20 | 0:40 - 2:40 | 1:12 - 1:35 |
| Time between request and arrival at SC (h:min) |  |  |  |
| All | 5:23 | 2:04 - 16:07 | 4:34 - 6:39 |
| Transfused | 3:09 | 1:14 - 13:41 | 1:37 - 11:52 |
| Not transfused | 5:34 | 2:13 - 16:10 | 4:50 - 7:03 |

IQR, interquartile range. TA, transfusional agency. SC, surgical center.

In 36 patients, the time between the request and the delivery of the blood sample in the TA was discrepant, ranging from 4h50min to 43 h. No outliers were identified for the time between the request and the patient’s admission to the SC. Figure 1 illustrates the time distribution between the request for the surgical reserve and its delivery to the TA and between the patient’s arrival at the SC.

**Figure 1.**
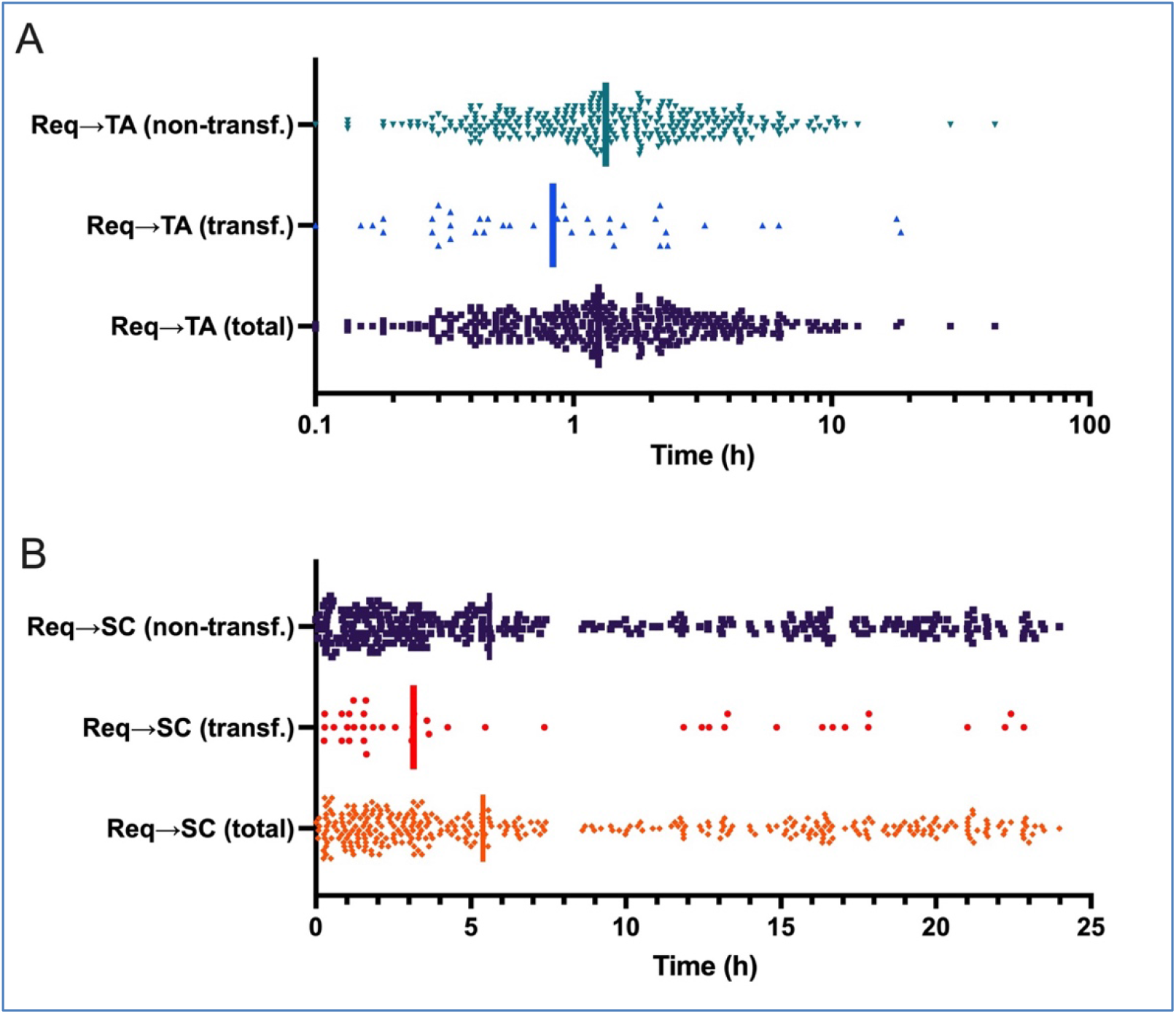
A, Distribution, in log_10_ scale, of times (in hours) from the request (Req) for blood concentrate reservation to the receivement at the transfusion agency (TA), according to the transfusion status (transfused, non-transfused, and total). B, Linear distribution of times from the request for blood concentrate reservation and the patient’s admission to the surgical center (SC). The vertical bars symbolize the medians.

There was no difference in the rate of transfusion in relation to sex (male n = 22; 11.6% vs. female n = 22; 13.1%; p = 0.66) or age (transf.: 59, 1 ± 22.3 years vs. non-transferred: 58.4 ± 22.2; p = 0.83). However, there was a higher percentage of transfusion in patients undergoing major surgery (major: n = 37, 14.8% vs. minor: n = 7, 6.5%; p = 0.027) and non-orthopedic surgery (ortho.: n = 9, 4.0% vs. non-ortho.: n = 35, 26.9%; p < 0.001).

## Discussion

This study observed a low probability of transfusion and a high C/T ratio in the sample. Still, there was a relative speed in the delivery of requests for blood concentrates, especially for patients who were effectively transfused. Most patients were referred to the SC within 12 h of the reservation request, and for patients effectively transfused, the median time was only three hours.

A small prevalence of males was observed in the sample (non-significant), consistent with their greater presence in emergencies (accidents and violence), and agrees with other studies^11,12^ that obtained similar results. The mean age observed was similar to that observed in the literature^11^ and was not different between transfused and non-transfused patients.

The reserve of blood for surgical procedures is an activity aimed at the safety of patients with the potential for bleeding between the intra- and immediate postoperative periods. Before performing any surgical intervention, even if considered urgent and emergency, the patient’s risk of bleeding is evaluated through clinical characteristics, laboratory parameters, and particularities of the surgery that will be performed. Due to these findings, it is requested to reserve blood or blood products that the patient will possibly need^13^.

However, this practice must be carried out consciously since it is an expensive activity with high costs for the health system. The transfusion of blood components involves costs from collecting blood units, transport, storage, preparation for transfusion or reservation within hospital units, and conducting pre-transfusion immunohematological tests so that the request cannot be carried out the wrong way^14,15^.

The request for reserve blood concentrates proved to be much greater than their actual use, following the high C/T ratio. The ratio between typed and transfused bags is an index used worldwide to measure the efficiency in the use of blood products. A study conducted by Zewdie et al.^8^ revealed an equally high rate (7.6), indicating an insignificant use of blood. Bhuthia et al.^16^, in turn, observed mostly lower C/T ratios (but ranging from 0.75 to 30.5), especially in oncological surgery. An Iranian study^17^ also observed a lower C/T (3.7), but in a sample essentially of elective surgery.

The probability of transfusion is another valuable tool to assess the efficiency in the requisition and use of blood components. It has also been used as an indicator, considered ideal if it is greater than 30%. In this study, a very low rate (12.3%) was observed, reflecting an excess of requests to reserve the service. Zewdie et al.^8^ and Yazdi et al.^17^ observed similar probability (15.3% and 16.8%, respectively), while other authors observed higher values^16^.

The TI observed in this study (0.27) was also below that considered adequate by the literature, indicating approximately one transfused bag for every four typed patients. As with other indexes, the variability of TI in different studies points to different efficiencies of each service. Shrestha et at.^18^ observed a TI of 1.09 bag/patient in a general hospital in Kathmandu, Nepal, with the worst rates observed in general surgery (0.88) and the best in pediatrics and dialysis (1.30). Alternatively, the study by Zewdie et al.^8^ also indicated an insufficient use of blood, with a TI of 0.29, while in a Nigerian study, a reasonable TI (0.9) was observed in orthopedic patients, whether elective or not, ranging from 0 to 2.619. An adequate TI of 0.77 was also observed in an Ethiopian tertiary hospital in surgical, obstetric, and gynecological cases^5^.

It is necessary to emphasize the importance of elaborating protocols to optimize the use of blood components and the work process involved in blood transfusion. This is considered an effective strategy for organizing and reducing the time and expense of unnecessary materials, aiming to reduce the number of transfusions without compromising the clinical outcome^15^.

This study investigated the time interval to making a reservation request and the time between the request and the patient’s arrival at the SC. A median time between the medical request and its delivery at the TA of 1h15min was observed, while between the request and the patient’s arrival at the SC was 5h23min. It is important to note that the median time interval was just over three hours among properly transfused patients. Despite being an important indicator to measure the efficiency of blood transfusion, few studies have addressed the response time of this workflow^20,21^, nor is there any standardization or threshold of acceptable times described in the literature.

The response time from the request chain to the transfusion was studied in an Indian tertiary hospital, where the median time between the request and its arrival at the blood bank was 47 min, the bag preparation time was 55 min, and the total time to transfusion was 135 min^22^. The authors considered this interval excessively long, mainly due to the delay in sending samples to the blood bank (34 min), shortages and messengers who completed the task, and the physical distance between sectors. Alternatively, a South Korean study observed that the institutional response time (up to 30 min) was achieved in 85.9% of blood requests, indicating another scale of efficiency^23^. Even shorter response times (8 to 12 min) were observed in two large North American teaching hospitals^6^.

However, it is essential to emphasize that the abovementioned studies do not refer to the request for a surgical reserve but to already indicated blood transfusions, with no margin for the interruption of the process. In the present study, it is worth noting that there was a considerable time gap between the request and the arrival of the transfused patient at the SC, indicating a reasonable working window for the nursing and medical staff. It was not possible to find any other national publication studying this temporal flow.

The administration of blood with urgency criteria is directed to maintain survival, but its use is usually preceded by the infusion of crystalloid or colloid solutions, both pre- and intraoperatively^14^. In most cases, there is adequate vital stabilization, and blood transfusion becomes a procedure with less urgency, and it is possible to wait for the determination of the ABO/Rh blood group and the performance of the compatibility tests^14,24^. For refractory cases or those with excessive volume loss, the transfusion of red blood cells from type O-negative donors without a compatibility test (emergency transfusion modality) is used. Although this situation is not directly linked to emergency surgeries or preoperative reservations, the adequacy of the service’s response time is essential for the maintenance of life. It is essential to mention that pre-transfusion exams take an average of 15 min to be performed, so the delays in the process are much more bureaucratic issues, such as taking and bringing samples, ordering, and picking up the released bag, among others.

Based on the premise that delays in blood supply can put the patient’s life at risk, the professionals involved in the transfusion act (doctors, nurses, nursing technicians, blood bank and nursing technicians) must understand the importance and seriousness involved in the entire transfusion process. It must be emphasized that any error, from inappropriate requests to the delivery of blood components in a longer time than expected, can cause irreversible damage to the patient. Therefore, all professionals involved must understand that the safety and efficacy of the transfusion process depend on them^25^.

Furthermore, implementing strategies to optimize blood component requisition is essential to reducing costs and waste^3^.The elaboration of time goals by the teams can be a decisive factor in improving the processes^26^. Mapping the structure of blood banks and transfusion agencies is another critical tool to leverage improvements. A study carried out in the state of Minas Gerais, involving the reference service (Hemominas), of which the hospital in this study is part, suggested adopting performance criteria for transfusion services and an external quality control program for the entire state^27^.

In addition to being observational and unicentric, within the limitations of this study, we can mention the absence of a data acquisition protocol defined in the medical record, with the need to search and consolidate the indicators from various forms and electronic databases. Still, it was impossible to measure the processing time of the packed red blood cells in the TA and the precise start of the transfusion, the latter due to the imprecision of the medical and nursing staff’s notes. It was also not feasible to define differences between the request times based on the shift in which the flow was started. We can predict that the response times for requests made at night and dawn will be longer than those made during the day due to the smaller number of workers and increased overload. Finally, it was impossible to compare the indicators of patients who had the reservation request made already in the SC, in an imminent nature of blood transfusion, due to the small number of cases.

## Conclusion

It was possible to analyze the rates of referral and request and the actual use of blood bags in patients undergoing urgent surgery in a teaching hospital. There was a significant discrepancy between the number of requests for blood supply and its effective use, given that a small portion of patients admitted to the SC of the investigated institution, considered urgent, actually received the planned transfusion. A long time was also observed between the request for the reservation and its referral to the TA, but this was mitigated by the high time between the request and the patient’s referral to the SC.

## Data Availability

All data produced in the present work are contained in the manuscript.

## Acknowledgements

We thank the pharmacist Mileni Rossi da Silva for providing access and guidance in collecting data from the Transfusion Agency.

## References

1. World Health Organization. WHO Guidelines for Safe Surgery 2009: Safe Surgery Saves Lives. WHO; 2009. Accessed November 22, 2021. https://www.who.int/teams/integrated-health-services/patient-safety/research/safe-surgery

2. Oliveira EM, Reis IA. What are the perspectives for blood donations and blood component transfusion worldwide? A systematic review of time series studies. Sao Paulo Med J. 2020;138(1):54–59. doi:10.1590/1516-3180.2019.0415.r1.06112019

3. Alves JL, Neto Clbg, Hékis HR, Contreras RC, Lins HWC, Melo ASP. Proposta de um novo protocolo de reservas de hemocomponentes para cirurgias em um hospital universitário de Recife-Pernambuco. Rev Bras Inov Tecnol Saúde. 2019:30. doi:10.18816/r-bits.v8i4.15647

4. Brasil, Ministério da Saúde, Agência Nacional de Vigilância Sanitária. Resolução RDC no 34, de 11 de junho de 2014. Dispõe sobre as Boas Práticas no Ciclo do Sangue. Published online 2011. Accessed December 14, 2021. https://saude.rs.gov.br/upload/arquivos/carga20170553/04145350-rdc-anvisa-34-2014.pdf

5. Belayneh T, Messele G, Abdissa Z, Tegene B. Blood Requisition and Utilization Practice in Surgical Patients at University of Gondar Hospital, Northwest Ethiopia. J Blood Transfus. 2013;2013:758910. doi:10.1155/2013/758910

6. McClain CM, Hughes J, Andrews JC, et al. Blood ordering from the operating room: turnaround time as a quality indicator. Transfusion. 2013;53(1):41–48. doi:10.1111/j.1537-2995.2012.03670.x

7. Isidoro REC, Silva KFN, Oliveira JF, Barichello E, Pires PS, Barbosa MH. Solicitação de reserva e preditores para hemotransfusão em cirurgias eletivas de fratura de fêmur. Texto Contexto Enferm. 2019;28:e20180129. doi:10.1590/1980-265x-tce-2018-0129

8. Zewdie K, Genetu A, Mekonnen Y, Worku T, Sahlu A, Gulilalt D. Efficiency of blood utilization in elective surgical patients. BMC Health Serv Res. 2019;19(1):804. doi:10.1186/s12913-019-4584-1

9. Marcondes SS, Carrareto AR, Zago-Gomes MP, Orletti MPSV, Novaes Aczl. Evaluation of the use of blood in surgeries as a tool to change patterns for requesting blood product reserves. Clinics. 2019;74:e652. doi:10.6061/clinics/2019/e652

10. Motulsky HJ, Brown RE. Detecting outliers when fitting data with nonlinear regression – a new method based on robust nonlinear regression and the false discovery rate. BMC Bioinformatics. 2006;7(1):123. doi:10.1186/1471-2105-7-123

11. Lobo SM, Vieira SR, Knibel MF, et al. Anemia e transfusões de concentrados de hemácias em pacientes graves nas UTI brasileiras (pelo FUNDO-AMIB). Rev Bras Ter Intens. 2006;18(3):234–241. doi:10.1590/s0103-507x2006000300004

12. Lima LP de, Menezes KP, Gadelha D de L, et al. Perfil de transfusão sanguínea e hemocomponentes em um hospital de urgência em Rio Branco. South Am J Basic Educ Tech Technolol. 2021;1(8):248–262.

13. Sekine L, Wirth LF, Faulhaber GAM, Seligman BGS. Análise do perfil de solicitações para transfusão de hemocomponentes no Hospital de Clínicas de Porto Alegre no ano de 2005. Rev Bras Hematol Hemot. 2008;30(3):208–212. doi:10.1590/s1516-84842008000300009

14. Neves MS de A, Delgado RB. Suporte hemoterápico ao paciente em emergência médica. Rev Med Minas Gerais. 2010;4(20):568–577. http://rmmg.org/exportar-pdf/338/v20n4a14.pdf

15. Bastos SL, Martins JCC, Oliveira ML, et al. Uso de hemocomponentes em hospital de médio porte em Belo Horizonte, Minas Gerais. Rev Médica Minas Gerais. 2014;Supl 6(24):S54–S60. doi:10.5935/2238-3182.20140086

16. Sg B, Srinivasan K, Ananthakrishnan N, Jayanthi S, Ravishankar M. Blood utilization in elective surgery - Requirements, ordering and transfusion practices. Nat Med J India. 1997;4(10):164–168.

17. Yazdi AP, Alipour M, Jahanbakhsh SS, Gharavifard M, Taghavi M. A Survey of Blood Request Versus Blood Utilization at a University Hospital in Iran. Archives Bone Jt Surg. 2016;4(1):75–79. doi:10.22038/abjs.2016.4710

18. Shrestha AN, Aryal BB, Poudel A, et al. Blood transfusion practices in a tertiary care hospital in Nepal. J Pathology Nepal. 2020;10(2):1728–1732. doi:10.3126/jpn.v10i2.30424

19. Adegboye MB, Kadir DM. Maximum surgical blood ordering schedule for common orthopedic surgical procedures in a tertiary hospital in North Central Nigeria. J Orthop Trauma Surg Rel Res. 2018;1(13):6–9.

20. Novis DA, Friedberg RC, Renner SW, Meier FA, Walsh MK. Operating Room Blood Delivery Turnaround Time. Arch Pathol Lab Med. 2002;126(8):909–914. doi:10.5858/2002-126-0909-orbdtt

21. Pati HP, Singh G. Turnaround Time (TAT): Difference in Concept for Laboratory and Clinician. Indian J Hematol Blo. 2014;30(2):81–84. doi:10.1007/s12288-012-0214-3

22. Agnihotri N, Agnihotri A. Turnaround Time for Red Blood Cell Transfusion in the Hospitalized Patient: A Single-Center “Blood Ordering, Requisitioning, Blood Bank, Issue (of Blood), and Transfusion Delay” Study. Indian J Crit Care Med. 2018;22(12):825–830. doi:10.4103/ijccm.ijccm_403_18

23. Lee AJ, Kim SG. Analysis of Turnaround Time for Intraoperative Red Blood Cell Issues: A Single-Center Study. Lab Med. 2017;48(3):277–281. doi:10.1093/labmed/lmx016

24. Como JJ, Dutton RP, Scalea TM, Edelman BB, Hess JR. Blood transfusion rates in the care of acute trauma. Transfusion. 2004;44(6):809–813. doi:10.1111/j.1537-2995.2004.03409.x

25. Silva EM, Vieira CA, Silva FO, Ferreira EV. Desafios da enfermagem diante das reações transfusionais. Rev Enferm UERJ. 2014;25(0):e11552. doi:10.12957/reuerj.2017.11552

26. Yazer MH, Deandrade DS, Triulzi DJ, Wisniewski MK, Waters JH. Electronic enhancements to blood ordering reduce component waste. Transfusion. 2016;56(3):564–570. doi:10.1111/trf.13399

27. Brener S, Carvalho RVF de, Ferreira ÂM, Silva MMF, Valle MCR de, Moraes-Souza H. Physical and operational infrastructure of transfusion services of the public blood bank network in the State of Minas Gerais, Brazil, 2007/2008. Rev Bras Hematol Hemoter. 2010;32(6):455–462. doi:10.1590/s1516-84842010000600009

